# DNA methylation-based age acceleration observed in IDH wild-type glioblastoma is associated with better outcome - including in elderly patients

**DOI:** 10.1101/2022.02.03.22270104

**Authors:** Pierre Bady, Christine Marosi, Michael Weller, Bjørn H. Grønberg, Henrik Schultz, Martin J. B. Taphoorn, Johanna M. M. Gijtenbeek, Martin J. van den Bent, Andreas von Deimling, Roger Stupp, Annika Malmström, Monika E. Hegi

## Abstract

Elderly patients represent a growing proportion of individuals with glioblastoma, who however, are often excluded from clinical trials owing to poor expected prognosis. Little is known about molecular features that would justify different treatment modalities, not related to frailty and co-morbidities.

We investigated the DNA methylome (450k) for age related associations using 4 different glioblastoma datasets (mean age, years: DKFZ, 60; TCGA, 62; Nordic, 71; EORTC-26981/NCIC-CE.3 & Lausanne-Pilot, 54), including two are cohorts from clinical trials. We excluded patient samples not compatible with glioblastoma grade 4 WHO-CNS-5 classification 2021. The combined methylome was interrogated for differences based on age, DNA methylation (DNAm) age acceleration (DNAm age “Horvath-clock” minus patient age), methylation-based classification (Heidelberg), entropy, and functional methylation of DNA damage response (DDR) genes.

Age dependent methylation included 19 CpGs (p-value≤0.1, Bonferroni corrected), comprising *ELOVL2* that is part of a 13-gene forensic age predictor. Three CpGs met our criteria of *functional* methylation defined as correlation of methylation with expression (r≤-0.3) of the corresponding gene. Most of the age related CpGs (n=16) were also associated with age acceleration that itself was associated with a large number of CpGs (n=50551). Over 70% age acceleration-associated CpGs (n=36348) overlapped with those associated with the methylation based tumor classification (n=170759). Functional methylation of few DDR genes was associated with age acceleration (n=8), tumor classification (n=12), or both (n=4), the latter including *MGMT*. DNAm age acceleration was significantly associated with better outcome in the trial cohorts of the EORTC/NCIC & Lausanne Pilot, and Nordic (treating elderly patients). Multivariate analysis included treatment (RT, RT/TMZ→TMZ; TMZ, RT), *MGMT* promoter methylation status, and interaction with treatment.

In conclusion, DNA methylation features of age acceleration are an integrative part of the methylation-based tumor classification (RTK I, RTK II, MES), while age seems hardly reflected in the glioblastoma DNA methylome.

## Introduction

Patients over the age of 70 represent an increasing fraction of individuals with glioblastoma (GBM) [38], yet eldery patients are often excluded from clinical trials due to the estimated poor prognosis and a short overall survival (6-9 months). Usually, a shorter duration radiation therapy schedule in association with temozolomide (TMZ) is favored for elderly patients aiming to shorten the treatment duration in view of the overall short life expectancy. Hypofractionated radiotherapy (15 × 2.66 Gy) has been shown clinically equivalent to standard 30 × 2 Gy fractionation [41], in combination with TMZ chemotherapy this regimen has been superior to hypofractionated radiotherapy alone [40]. In methylated tumors, exclusive or initial TMZ chemotherapy alone may be an alternative to radiotherapy. Median overall survival remains unsatisfactory with treatment regimens. The choice of the treatment regimen depends on the patient’s biological rather than chronological age and frailty, and the tumor *MGMT* promoter methylation status [55]. Yet, a fraction of elderly patients have a more favorable outcome with disease control beyond 18-24 months. Little is known about the molecular make-up of GBM in older patients (7th and 8th decade of life) and whether it differs from a “general” glioblastoma IDHwt population. We have reported previously from the Nordic trial, treating newly diagnosed glioblastoma patients aged 60 or older that the *MGMT* promoter methylation status is not associated with patient age [31]. The predictive value of the *MGMT* status for benefit from TMZ is supported by the aforementioned trials. Expectedly, mutations in the isocitrate dehydrogenase gene 1 (*IDH1*), a hallmark of gliomas in young adults, are rare in the elderly [31, 56]. Moreover, no obvious GBM-specific genetic alterations have been associated with the age of the patients, once removing IDHmut cases that are recognized and classified as a distinct disease entity since 2016 [29]. Hence, systematic molecular characterization is required, in order to justify distinct treatment modalities not owing to frailty of the patients, or suggesting new, more adapted (targeted) approaches that can be tested in clinical trials in this elderly patient population.

In this translational research study we set out to investigate the DNA methylome of adult IDH wild-type glioblastoma for age related differences. We used four data sets, including cohorts of patients treated for newly diagnosed GBM in clinical trials [8, 9, 31, 46, 47], whereof one recruited elderly patients [31]. The DNA methylome of cells is known to contain age related information reflected in age associated epigenetic changes. These are thought to arise from innate biological mechanisms, such as deterioration of epigenetic maintenance resulting in changes of DNA methylation over time that allowed the construction of accurate predictors of chronologic age. Such predictors are known as *epigenetic clocks* or *DNA methylation clocks* [12, 16, 20, 21, 58]. Furthermore, certain biological features or environmental factors have been associated with an *acceleration* of the DNA methylation age (DNAm age) in the blood, certain tissues, and cancer. This difference between chronologic age and estimated DNAm age is called DNAm age acceleration, and has been found associated with obesity, and smoking, but also Down syndrome or cancer, as reviewed by Horvath and Raj [22]. It has been proposed that biological age acceleration may hold the potential for disease specific biomarkers or frailty measures for individuals [5, 22, 32]. In addition, the tumor DNA methylome comprises information on cell of origin, plus tumor development related alterations that allow accurate tumor classification and identification of new tumor entities [9, 35]. Previous studies have reported on associations of tumor DNAm age with chronologic age in adult glioma and GBM [28, 61]. However, these studies included glioma of variable grades and subtypes, including IDH mutant (IDHmt) glioma that are epigenetically and clinically a distinct entity [10]. In particular, IDHmt gliomas exert a glioma CpG island methylator phenotype (G-CIMP) and mostly affect young adults [37, 49]. These tumors have a different etiology and epigenetic and genetic make-up that is associated with better prognosis, and are classified as a different tumor entity since the WHO 2016 update [29]. We have thus excluded all IDHmt or G-CIMP+ tumors and as well as tumors reclassified as “other” gliomas according to the recent update of the WHO CNS5 classification 2021 [30] in all datasets used for this study. Only tumors that meet the criteria for glioblastoma according to the most recent 2021 WHO classification have been considered for this analysis.

## Material and methods

### Patient samples

Two sets of newly diagnosed GBM samples were obtained from patients treated in clinical trials according to pre-specified clinical criteria (Nordic, n=116; EORTC 26981/NCIC CE.3 pooled with the Lausanne Pilot trial, n=219). The constituted cohorts were restricted to patients for whom enough frozen or paraffin embedded tissue was available, thus excluding cases with diagnostic biopsies only. The cohorts overlaps largely with those for which *MGMT* promoter methylation status was reported from the original trials [18, 19, 31]. The Nordic phase III trial randomized patients 60 years or older and a WHO performance of 3 or less (if it was due to neurological deficit, but the general condition of the patient needed to be PS 0-2) [54] to one of two regimens of RT (60 Gy in 30 fractions, or 34 G in 10 fractions) or standard dose TMZ (200mg/m^2^, days 1-5 every 28 days) [31] (trial registration number ISRCTN81470623). The Lausanne Pilot (LN-Pilot) trial and the EORTC 26981/NCIC CE.3 trial (trial registration number NCT00006353) recruited patients between the ages of 18 and 70, and a WHO performance of < 2 [46, 48]. Patients in the uncontrolled phase II LN-Pilot trial received the TMZ/RT→TMZ standard regimen (the current standard of care), and patients in the pivotal trial EORTC 26981/NCIC CE.3 were randomized to RT (60 Gy in 30 fractions) or TMZ/RT→TMZ. Patients signed consent for translational research according to institutional and international guidelines and regulations and was conducted in accordance with the Declaration of Helsinki. See detailed Consent and Ethics declaration at the end of the manuscript.

### DNA methylation analysis

For genome-wide DNA methylation analysis DNA was isolated from macro-dissected tumor tissue (frozen samples: DNeasy Blood & Tissue Kit, Qiagen; formalin fixed paraffin embedded (FFPE) samples: EX-WAX™ Paraffin-embedded DNA Extraction Kit, S4530; Merck KGAa) and quantified (Quant-iT™ PicoGreen® dsDNA Assay Kit, #P7589, Life Technologies). DNA samples were analyzed on the Human Methylation 450K BeadChip (Illumina, San Diego CA, USA) at the Genomics platform of the University of Geneva. FFPE-derived tumor DNA samples were processed after passing a PCR-based quality control (Infinium HD FFPE QC Assay Protocol). DNA samples were subjected to bisulfite treatment (EZ DNA Methylation-Gold™ Kit, Zymo Research) as previously described, and FFPE samples were analyzed in separate batches after pretreatment with the restauration kit as recommended (llumina) [3, 24].

### Data availability

The datasets are available in GEO (http://www.ncbi.nlm.nih.gov/geo/) under the accession numbers GSE195684 for the Nordic trial samples, and the samples from the LN-Pilot trial and the EORTC 26981/NCIC CE.3 trial are available at GSE195640, or GSE60274. The latter comprises data from a subset of GBM samples of the EORTC-NCIC & Pilot trials and 5 non-tumoral brain tissue samples that have been previously published [24]. Methylation data from an additional 5 non-tumoral brain tissues used, is available under GSE104293 [3].

### External datasets

External datasets comprised the GBM dataset from The Cancer Genome Atlas for which RNA-seq and HM-450k data are available and using corresponding annotations [8] (TCGA; n =113; dbGaP accession number phs000178.v9.p8; http://cancergenome.nih.gov), and a GBM set with HM-450k data from the DKFZ [9] (n=235; GEO accession number GSE109381).

### DNA methylation preprocessing

The CpG probes with detection p-values > 0.01, located on the sex chromosomes, or in SNPs were removed. The functional normalization [13] for Illumina 450k arrays includes noob (normal-exponential out-of-band) background correction, dye-correction (chemistry I vs II) and RUV-2 step (removing unwanted variation) based on control probes. This normalization was performed by the function **preprocessFunnorm** from the R package **minfi**. DNA methylation was summarized by M values [11]. The ComBat procedure [23] was used to aggregate the four datasets to limit experimental variation and batch effects across the four datasets.

### Additional metrics based on DNA methylation

The DNA methylation status of the *MGMT* promoter and the *MGMT* score (logit-transformed probability) were determined using the MGMT-STP27 regression model based on HM-450k data [2, 4]. The tumor purity (**HMpurity**) of each sample was estimated as previously proposed [3] using the GBM TCGA datasets.

### Molecular Subtypes

The G-CIMP status was determined by unsupervised clustering (Ward’s algorithm with Euclidean distance) as previously reported [3] and served as approximation for the IDH mutation status, as this information was not available for all samples in any of the datasets. All G-CIMP positive samples were removed. Molecular subtypes were obtained using the classification procedure of central nervous system tumors based on the analysis of DNA methylation patterns [9] (version v11b4, www.molecularneuropathology.org). For the main analyses of this study, we considered only glioblastoma classified as mesenchymal (GBM_MES), RTK I (GBM_RTK_I) or RTK II (GBM_RTK_II).

### DNAage and age acceleration

DNA methylation-based estimates of age (DNAm age) were calculated using ElasticNet regression [62] using 353 CpG sites selected by Horvath clock [20, 22]. The DNA methylation data were calibrated before the computation of DNAm age as recommended [20]. The metric called Age Acceleration (**Accel**) was obtained by subtraction of chronological age (**age**) from DNA methylation age (**DNAm age**). For subsequent analysis, 306 clock probes were conserved after filtering and aggregation of the four datasets (detection p-values > 0.01).

### DNA methylation Entropy (HME)

Estimation of the DNA methylation entropy (HME) is given by the normalized Shannon entropy [44] adapted for methylation fraction (p) given by Beta values [16]. For the i^th^ methylation marker, two states were possible: unmethylated (*1-pi*) or methylated (*pi)* and the maximal entropy is given by log(2). The DNA methylation Entropy (HME) for N methylation markers is computed as follow:

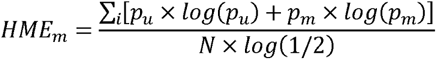

The HME metrics were defined for all CpGs (global HM-entropy) and for the 12 strata constituted by the Island regions (CpG islands, shores, shelves or open sea) and promoter location status (promoter or not in promoter). The HME table was described by variation partitioning [7] for age, age acceleration and GBM classification. The results are illustrated in a Venn diagram containing the variation fractions for the three supplementary variables.

### Expression

Gene expression from RNA sequencing (RNA-seq) data (Level 3), from the TCGA GBM dataset selected for this study, was quantified for the transcript models using RSEM [27] and normalized within samples to a fixed upper quartile for TCGA. Further details are available at the DCC data portal of TCGA. Gene-level data were restricted to genes expressed in at least 70% of samples. The complete dataset was normalized by the VOOM procedure [26].

### Statistical Analysis

#### Detection of DMP

The associations of CpG-probes with age or age acceleration were investigated by model comparison based on linear models with F test. The Bonferroni procedure was used to account for multiple testing comparisons. A differentially methylated position (DMP) was defined as a candidate for which the q-value was less than 0.1.

#### Detection of functional methylation

The correlation of methylation with the expression level for each CpG-probe located in a gene promoter within 1500 nucleotides up- or down-stream of the transcription start site (TSS) was estimated by Spearman correlation test. A methylated position was defined as functional when the q-value was less than 0.05 and the correlation coefficient was inferior to -0.3 (negative effect on gene expression). The gene locations were based on Homo sapiens data from UCSC build hg19.

#### Cox regression model and other tests

For the continuous variables Wilcoxon test (t) or Kruskall and Wallis test (a) were used to test the differences between two or more groups. The independence between qualitative variables and groups was tested with Pearson’s Chi-squared with Yates’ continuity correction. Survival univariate and multivariate models were computed by Cox proportional hazards regression model [51]. The association tests with GBM classification, study origins and the interaction between these both variables were performed by model comparison using Wald’s test with sandwich estimation of the covariance matrix and F-statistic. The covariance of the models is estimated by sandwich methods with the type version *HC3* [60] to compensate heteroscedasticity. Principal Component analyses and the permutation multivariate analyses of variance (ADONIS) [1] using Euclidean distances were used to investigate the association between additional variables (e.g. age, age acceleration or GBM classification) and DNA methylation data. Analyses and graphical representations were performed using **R-3.3.2** and the R package rms and survival (R2014, rms2014 and survival 2014).

## Results

### Patient characteristics

To investigate age dependent DNA methylation features of GBM, we included patients treated for newly diagnosed GBM in clinical trials; the Nordic trial (n=116) treating elderly patients, and the EORTC 26981/NCIC CE/3 trial, including patients from the preceding phase II study, the Lausanne Pilot (LN-Pilot) trial (n=219). The DNA methylome was established on the 450K platform. The baseline description of the full cohorts is presented in Supplementary Table S1. The DNA methylation-based tumor classification [9] and its association with age is visualized in Supplementary Figure S1. Of note, the patient cohort constituted of the EORTC/NCIC & LN-Pilot trials exhibited more diversity in glioma subtypes than the Nordic cohort.

Additional two external GBM datasets were included in the analyses. The age ranges of the patients in the four full GBM datasets, after removing all IDH mutant /G-CIMP+ samples, were as follows: Nordic (n=115) 60 to 83 years (median 70, SD 4.8 years), EORTC/NCIC & LN-Pilot (n=195) 26 to 70 years (median 55, SD 9.39 years), TCGA (n=113) 21 to 85 years (median 62, SD 11.9 years), and DKFZ (n=235) 18 to 86 years (median 59, SD 13.3 years).The tumors of the four datasets were classified according to the Heidelberg DNA methylation-based classifier [9]. Most GBM were classified as MES, RTK I or RTK II (88% for TCGA, 91% for the other 3 datasets), with few samples belonging to GBM-MID or GBM-G34 (no GBM-G34 in Nordic, Supplementary Figure S1) or others that are now considered distinct tumor entities in the updated WHO classification 2021, and few non-GBM classifications. This study was subsequently restricted to IDHwt GBM subtypes MES, and RTK I or II, for which the baseline characteristics of the patients are summarized in Table 1 for each dataset.

**Table 1:** Baseline description and parameters of GBM datasets

| **\*Datasets** | **DKFZ** | **EORTC/NCIC & LN-Pilot** | **Nordic** | **TCGA** |
| --- | --- | --- | --- | --- |
| **N** | 214 | 177 | 105 | 99 |
| **Sex: P = 0.1994** |  |  |  |  |
| F | 101 (47) | 66 (37) | 40 (38) | 42 (42) |
| M | 113 (53) | 111 (63) | 65 (62) | 57 (58) |
| **Age (yrs): P < 0.0001** |  |  |  |  |
| min | 29 | 27 | 60 | 33 |
| max | 86 | 70 | 83 | 85 |
| mean (sd) | 60.34 ± 11.25 | 54.35 ± 8.98 | 70.82 ± 4.66 | 61.95 ± 10.64 |
| **DNAm age (yrs): P = 0.0001** |  |  |  |  |
| min | 40.02 | 39.56 | 57.90 | 30.11 |
| max | 174.01 | 167.42 | 180.10 | 186.71 |
| mean (sd) | 94.65 ± 22.58 | 94.13 ± 26.13 | 107.75 ± 25.17 | 98.71 ± 27.72 |
| **Age Acc (yrs): P = 0.1737** |  |  |  |  |
| min | -13.45 | -9.50 | -7.19 | -14.89 |
| max | 135.01 | 104.14 | 108.49 | 120.71 |
| mean (sd) | 34.31 ± 21.25 | 39.78 ± 24.71 | 36.93 ± 25.09 | 36.76 ± 26.67 |
| **GBM subgroup: P = 0.0141** |  |  |  |  |
| GBM_MES | 48 (22) | 66 (37) | 43 (41) | 31 (31) |
| GBM_RTK_I | 51 (24) | 30 (17) | 20 (19) | 19 (19) |
| GBM_RTK_II | 115 (54) | 81 (46) | 42 (40) | 49 (49) |
| **MGMT-STP27: P = 0.4871** |  |  |  |  |
| U | 100 (47) | 91 (51) | 55 (52) | 55 (56) |
| M | 114 (53) | 86 (49) | 50 (48) | 44 (44) |
| **MGMT score: P = 0.3412** |  |  |  |  |
| min | -5.58 | -10.48 | -4.09 | -11.68 |
| max | 6.98 | 7.35 | 8.16 | 6.34 |
| mean (sd) | 0.62 ± 3.59 | 0.22 ± 3.38 | 0.32 ± 3.31 | -0.23 ± 3.85 |
| **HM purity: P = 0.6177** |  |  |  |  |
| min | 0.31 | 0.42 | 0.42 | 0.39 |
| max | 0.99 | 0.95 | 0.98 | 0.98 |
| mean (sd) | 0.73 ± 0.14 | 0.74 ± 0.14 | 0.75 ± 0.14 | 0.75 ± 0.14 |
| **Global HM entropy: P = 0.9938** |  |  |  |  |
| min | 0.45 | 0.47 | 0.47 | 0.49 |
| max | 0.64 | 0.64 | 0.62 | 0.63 |
| mean (sd) | 0.55 ± 0.03 | 0.55 ± 0.03 | 0.55 ± 0.03 | 0.55 ± 0.03 |
| **HME Prom CpG: P = 0.2526** |  |  |  |  |
| min | 0.27 | 0.29 | 0.29 | 0.28 |
| max | 0.48 | 0.47 | 0.45 | 0.41 |
| mean (sd) | 0.36 ± 0.03 | 0.36 ± 0.03 | 0.36 ± 0.03 | 0.36 ± 0.03 |

\*datasets include GBM\_MES, GBM\_RTK I and GBM\_RTK II, only

The distribution of the tumors in the three GBM subclasses was significantly different between the four datasets (p=0.014, chi squared). There were less MES GBM comprised in the DKFZ dataset as compared to the others (22% less), likely due differences in patient selection of the study. There were no differences in the proportion of female and male patients, or the frequency of *MGMT* promoter methylation between the datasets.

After initial filtering 361745 CpGs were retained for subsequent analyses. After batch correction no effect of the four datasets was observed on the DNA methylation based organization of the samples as illustrated in a PCA (R2=0.002, p=1.00, Figure 1B). In contrast, the global organization of DNA methylation was significantly affected by GBM methylation subclasses (R2=0.083, p=0.01, Figure 1C). Similarly, no differences were observed for sample purity between the data sets (Table 1), but between the GBM subclasses (Wald’s test, p < 0.001, Table 2; Figure 1E).

**Table 2:** Wald tests for models with GBM classification and study origin, and interaction.

| Characteristic | MES | RTK I | RTK II | GBM classification |  | Study |  | GBM Class x Study |  |
| --- | --- | --- | --- | --- | --- | --- | --- | --- | --- |
|  | mean (sd) | mean (sd) | mean (sd) | *F-value | Pr(>F) | *F-value | Pr(>F) | *F-value | Pr(>F) |
| Age [yrs] | 60.175 (10.853) | 61.075 (11.927) | 60.834 (10.83) | 0.216 | 0.806 | 52.158 | <0.001 | 0.747 | 0.612 |
| DNAm age [yrs] | 89.246 (21.746) | 94.538 (24.999) | 104.106 (26.163) | 10.717 | <0.001 | 4.670 | 0.003 | 1.896 | 0.079 |
| Age accel [yrs] | 29.072 (19.566) | 33.463 (21.956) | 43.273 (25.652) | 10.480 | <0.001 | 1.457 | 0.225 | 2.456 | 0.024 |
| HM-purity | 0.592 (0.104) | 0.817 (0.105) | 0.802 (0.099) | 68.162 | <0.001 | 2.288 | 0.078 | 1.086 | 0.369 |
| Global HME | 0.556 (0.029) | 0.571 (0.027) | 0.545 (0.029) | 18.557 | <0.001 | 0.029 | 0.994 | 0.394 | 0.883 |
| HME prom CpG | 0.362 (0.032) | 0.361 (0.03) | 0.363 (0.029) | 0.086 | 0.917 | 0.147 | 0.932 | 1.815 | 0.094 |

\*F-values are computed with covariance matrix, estimated by sandwich estimation (type HC3) to reduce the effect of the variance heterogeneity.

**Figure 1.**
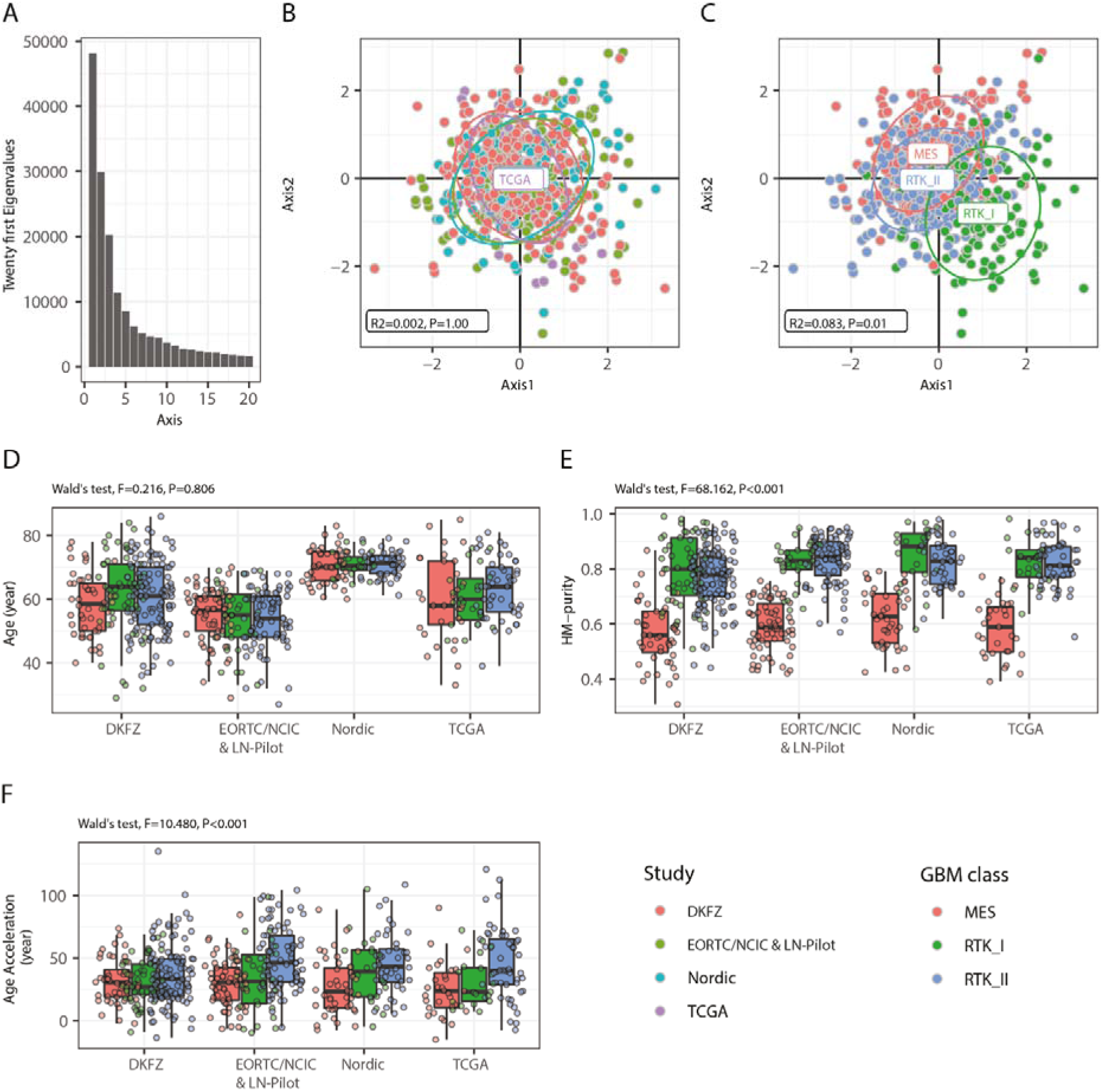
Organization of DNA methylation by dataset and GBM subtype. The representation of the inertia structure (A) shows three first distinct eigenvalues. The patient samples are represented on the first vectorial plane of the PCA of DNA methylation, annotated by study origin (DKFZ, red; EORTC & LN-Pilot, green; Nordic, blue; TCGA, pink; due to the overlap of the datasets, only one label is visible) (B) and methylation-based GBM classification, MES, red; RTK I, green; and RTK II, blue (C). The R-squared (R2) and p-value (99 permutations, ADONIS) testing the effect between the DNA methylation data and dataset (B) and GBM subtype (C) are indicated. The boxplot representations for age (D), sample purity (E), and DNAm age acceleration (F) are drawn, stratified by methylation-based GBM classification and study origin.

The lowest purity was associated with the MES GBM subtype (Figure 1E), in line with a more pronounced fraction of tumor infiltrating cells that has been associated with this subtype [53]. The Nordic trial, recruiting only elderly patients (Table 1), introduced a significant difference of age among the four datasets (Wald’s Test, p < 0.001, Table 2).

However, no age related association with the three GBM subtypes was observed (Wald’s Test, p=0.805, Table 2, Figure 1D). Finally, no association was observed between the GBM subtype and the WHO performance score at study entry (PS, scale 0 to 4, with higher values indicating greater disability [54]) (Cochran-Mantel-Henszel chi-squared test with stratification by study origin, p=0.720). These data was available for the patients treated in the EORTC 26981/NCIC CE/3, the LN-Pilot study (PS 0-2) and the Nordic trial (PS 0-3), respectively (Supplementary, Table S1).

### Age related differential methylation (DMP)

First, we analyzed the DNA methylation data for associations with the patients’ age. Age dependent methylation identified 19 CpGs (p-value ≤ 0.1, after Bonferroni correction; Supplementary Table S2). Of these, *ELOVL Fatty Acid Elongase 2* (*ELOVL2)* methylation at cg16867657 has previously been published as a biomarker for chronologic age (r=0.92) [14] and is part of forensic age predictors [36, 59]. Similarly, methylation levels of *Tripartite Motif Containing 59 (TRIM59)* have been associated with chronologic age [32, 59]. Several of the probes were associated with cancer relevant genes. Three CpGs met our criteria of *functional* methylation that we defined as negative correlation of methylation with expression (≤-0.3 and p-value adjusted for multiple testing ≤0.1) of the corresponding, annotated gene. These comprised functional CpG probes for *TRIM59, Twist Family BHLH Transcription Factor 1 (TWIST1)*, and *Nuclear Receptor Interacting Protein 3 (NRIP3)*, respectively (Supplementary Table S2). Functional methylation information was derived from the TCGA dataset that comprises RNA-sequencing data.

### DNA methylation age acceleration

Next, we determined DNAm age acceleration of the tumors that is defined as the DNAm age of the tumor, minus the patient’s age. The association of the patients’ chronologic age and the tumor DNAm age, determined with the Horvath clock, was modest, as illustrated in scatterplots for the datasets, EORTC/NCIC & LN-Pilot (r=0.322), TCGA (r=0.294) and the DKFZ dataset (r=0.400), (Figure 2), and weak for the Nordic dataset (r= 0.155). The small age range of this older population, may explain the latter (Table 1).

**Figure 2.**
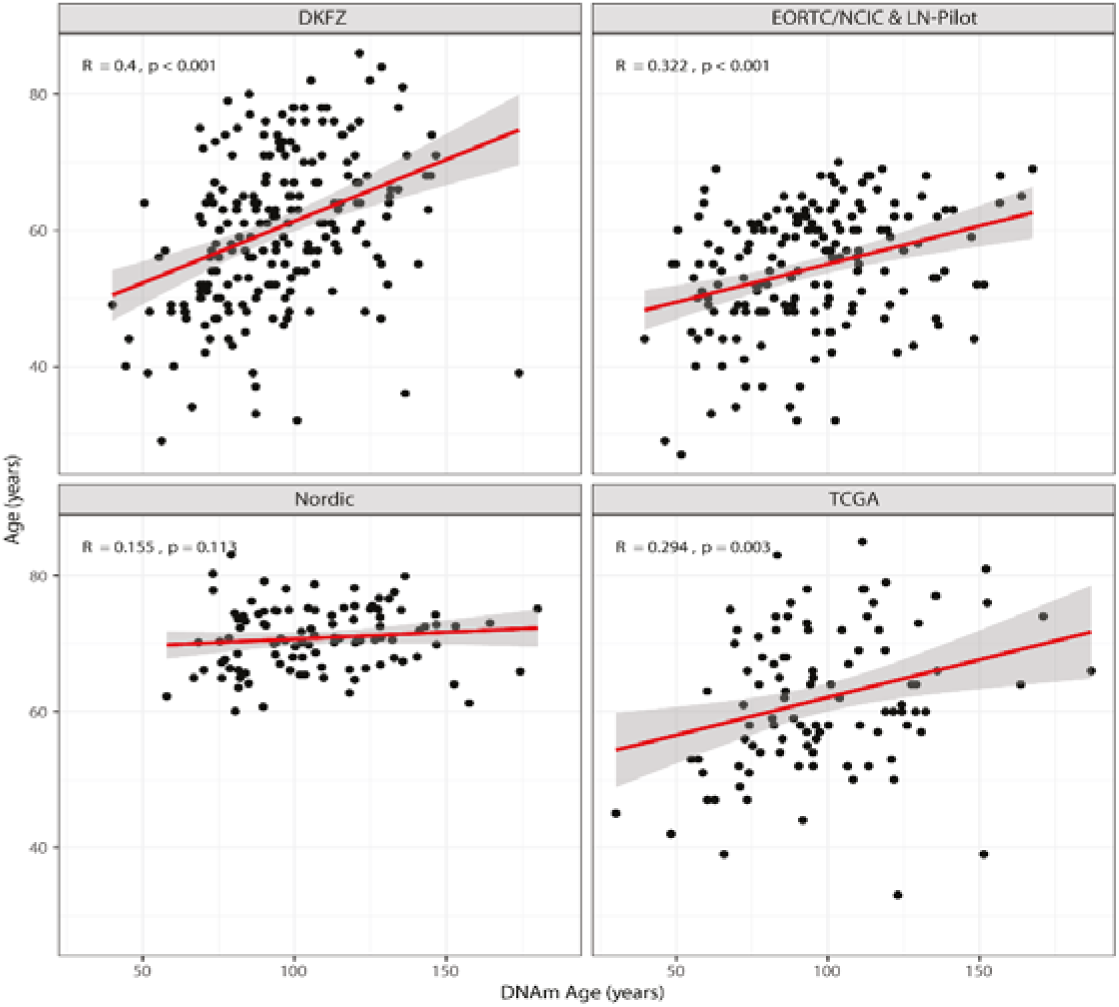
Chronological age versus DNAm age of GBM. Chronological age (observed age) versus DNAm age predicted by DNA methylation data is shown for the four datasets. The accuracy of the models is given by the Spearman’s coefficient correlation and regression model between observed and predicted age. The correlation values (|r| ≤ 0.4) show strong deviation between chronological age versus DNAm age of the GBM in the four studies.

The age acceleration was significantly different to zero (t test, p <0.001) with an averaged acceleration of 36.81 years and a standard deviation of 23.99 years. No differences in DNAm age acceleration were observed between male and female patients (p = 0.774, ANOVA, stratified by study). It is of note that the association between the numerous copy number variations (CNVs), characteristic for GBM, and the interrogated clock probes and DNAm age was weak. Only 7% of the variation of the DNAm age (p < 0.001) was explained, based on the regression of the four first axes from PCA of the clock probes, hence, excluding a strong confounding influence by CNVs.

A large number of CpGs associated with age acceleration were identified (DMP age accel, n=50551). Most of the age related CpGs (16 of 19) were also associated with age acceleration. By construction, age acceleration is not associated with the patients’ age. Interestingly, over 70% age acceleration associated CpGs (n= 36348) overlapped with those associated with tumor classification (n=170759) into the three methylation-based GBM subtypes (MES, RTK I, RTK II) (Figure 3A). Hence, it was not surprising that age acceleration was significantly associated with the GBM subtype (Wald’s test, p < 0.001, Table 2; Figure 1F). The tumors classified as GBM RTK II, trended to exhibit higher age acceleration than the two other GBM methylation subclasses. However, the sample purity constituted a weak confounding factor, with a Spearman correlation coefficient between DMAm age acceleration and purity of r = 0.290, (p < 0.001).

**Figure 3.**
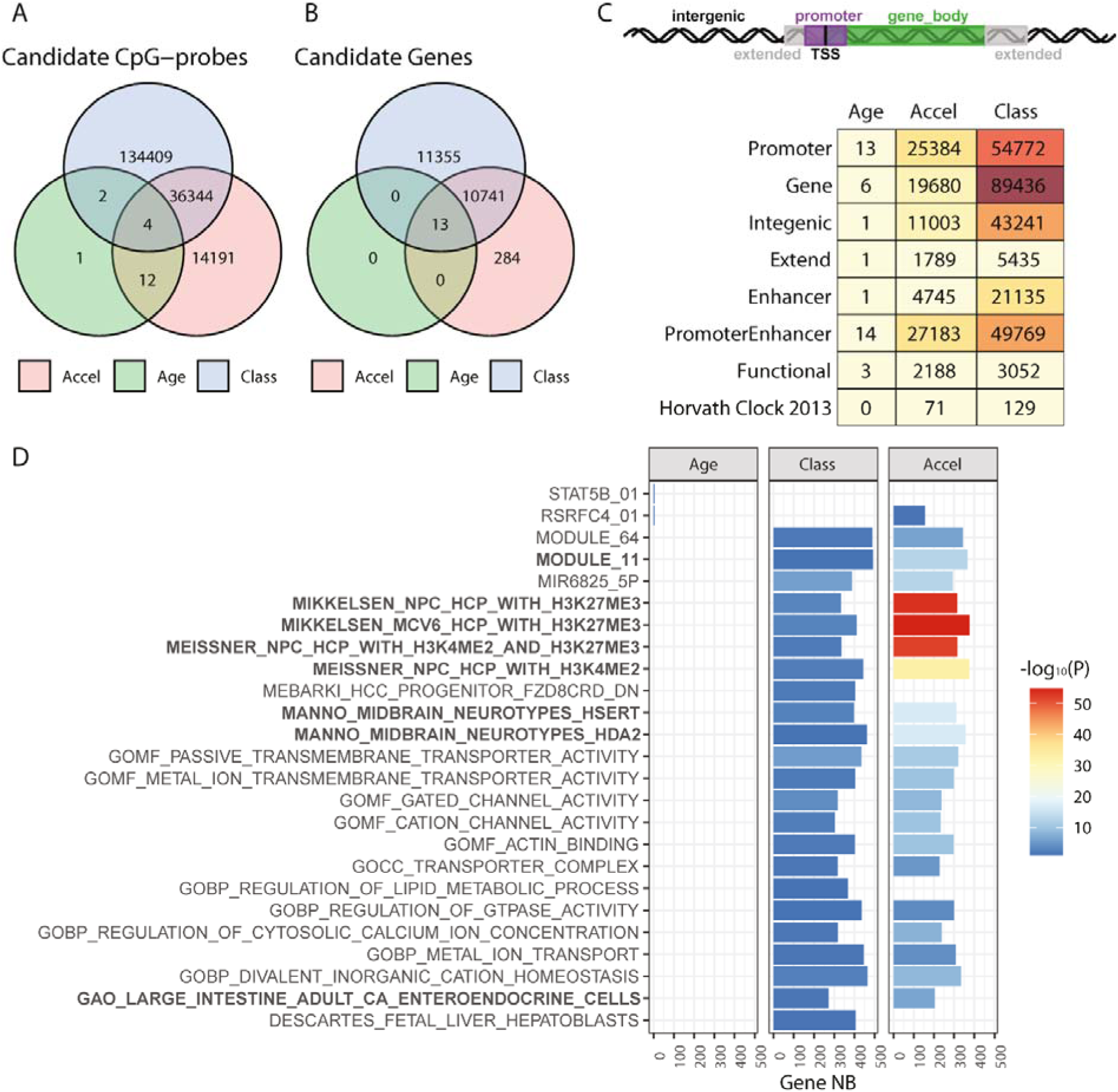
Associations of DNA methylome with age, DNAm age acceleration, and GBM subclassification. Methylome-wide association studies of the samples were performed for age (green), DNAm age acceleration (pink) and GBM classification (blue) and visualized in a Venn-diagram, for the interaction between the three sets of candidate CpG-probes (A) and the associated genes (B). The genomic location of the selected CpGs, their association with age, DNAm age acceleration, GBM classification, the CpG functionality, and association with the Horavth clock are summarized in (C). Gene set enrichment analysis (GSEA), using the MSigDB database, was established for candidate CpG-probes related to age, DNAm age acceleration and GBM classification. The list of gene sets (D) corresponds to the pathways significantly enriched for age and GBM classification. The number (NB) of genes per gene set is indicated and the p-value is represented by the color code. The gene lists overlapping with those associated with functional methylation (Additional File 1, Figure S2) are highlighted in bold.

The overlap of the CpGs fulfilling all three criteria, age, DNAm age acceleration, and methylation-based subclassification, comprised 4 CpGs that also included the probe in the *ELOVL2* promoter (Supplementary Table S2). This suggests that DNA methylation features of DNAm age acceleration are an integrative part of methylation-based tumor classification, while age seems only slightly reflected in the tumor DNA methylome. The breakdown of the selected CpGs by genome regions and function (e.g. promoter, gene body, enhancer, etc); the three variables, age, DNAm age acceleration and classification, and affiliation with the Horvath clock, is detailed in Fig 3C. It is of note that the observed significant association of 129 clock probes with the GBM classification was not dependent on their location on the 22 autosomes (Fisher’s test with p-value estimated by Monté-Carlo simulation, p=0.558).

### Pathway analysis of DNAm age acceleration and tumor classification

Next, we were interested in the pathways to which the genes belonged that were associated with DNAm age acceleration or classification. Performing signature analysis using MiSigDB (molecular signatures database; gene set enrichment analysis [GSEA], adjusted p≤0.1) and the CpGs associated with DNAm age acceleration yielded 1220 pathways. The top pathways were dominated by gene-sets characterizing epigenetic properties of neural precursor cells, or gene sets for midbrain neurotypes, and other progenitor cells, and axon development (Figure 3D) invoking developmental features and cell of origin. The tumor classification associated CpGs yielded 23 pathways, of which most (n=20) overlapped with those from DNAm age acceleration, in line with the large overlap of CpGs between the two criteria.

The common pathways were also dominated by gene sets characterizing epigenetic features of neural progenitors, included sets for transmembrane transporters and channels, and cancer gene sets (Module 11 and 64) (Figure 3D). The few CpGs associated with age were linked with gene sets of downstream targets of STAT5B (Signal Transducer And Activator Of Transcription 5) and RSRFC4 (alias of MEF2A, Myocyte-Specific Enhancer Factor 2A), with 3 of the 4 genes overlapping in the two sets, the latter was also associated with DNAm age acceleration.

Subsequently we looked only at the functional CpGs (Figure 3C), as they may shed light on associated biological mechanisms, and determined the associated pathways (GSEA, adjusted p≤0.1). The “functional” pathways associated with DNAm age accel (n=167) mostly (119, 71%) overlapped with those associated with classification (n=294) (Suppl Fig. S2). The overlapping “functional” pathways were dominated by gene-signatures characterizing epigenetic properties of neural precursor cells, or gene sets for midbrain neurotypes, and other progenitor cells, and developmental genes. In addition, signatures of immune cells, and some cancer related signatures were comprised, the latter including the Verhaak expression signature for mesenchymal GBM (Supplementary Figure S2). Interestingly, but not surprising, among the top 30 functional pathways associated with the methylation-based classification comprised three signatures of the expression based GBM classifier defined by Verhaak [52] (signatures for mesenchymal, classical, and proneural GBM). These observations support the overall consistency and biological relevance of the findings.

### Human Methylation Entropy (HME) and age

Subsequently, we investigated the genome-wide variation of DNA methylation that may affect regulatory functions and genomic/epigenomic stability of the tumors. For this purpose, we used the measure of the Human Methylation Entropy (HME) that quantifies the methylation complexity for a given CpG or a given genomic region [16]. Low heterogeneity/high similarity corresponds to low entropy scores (range 0 to 1; e.g. 100% methylation, 0 entropy; 50% methylation, organized in fully methylated and fully unmethylated alleles, 0.25 entropy; 50% methylation, organized randomly, 0.65 entropy, for more details see [43]). The entropy at 12 distinct genomic regions (promoter, shore, etc.) tested, showed characteristic features by region, with strikingly lower entropy levels within CpG promoter islands (Figure 4). This may suggest that CpG promoter islands present more homogenous DNA methylation patterns, e.g. methylated or unmethylated states, likely due to their direct regulatory function in gene expression. HME at different genomic regions (Supplementary Table S3) and global HME were associated with GBM subtype (Wald’s Test, p < 0.001, Table 2, Figure 4E). Indeed, HME was highest in RTK I for most genomic regions. In contrast, no difference between tumor subtypes was detected for HME in the promoter CpGs Islands (Wald’s Test, p = 0.917, Table 2, Figure 4F). No differences were observed between datasets at the distinct regions and over all regions combined. Finally, we performed variation partitioning of HM-entropy metrics to evaluate the contribution of age, age acceleration and GBM classification.

**Figure 4.**
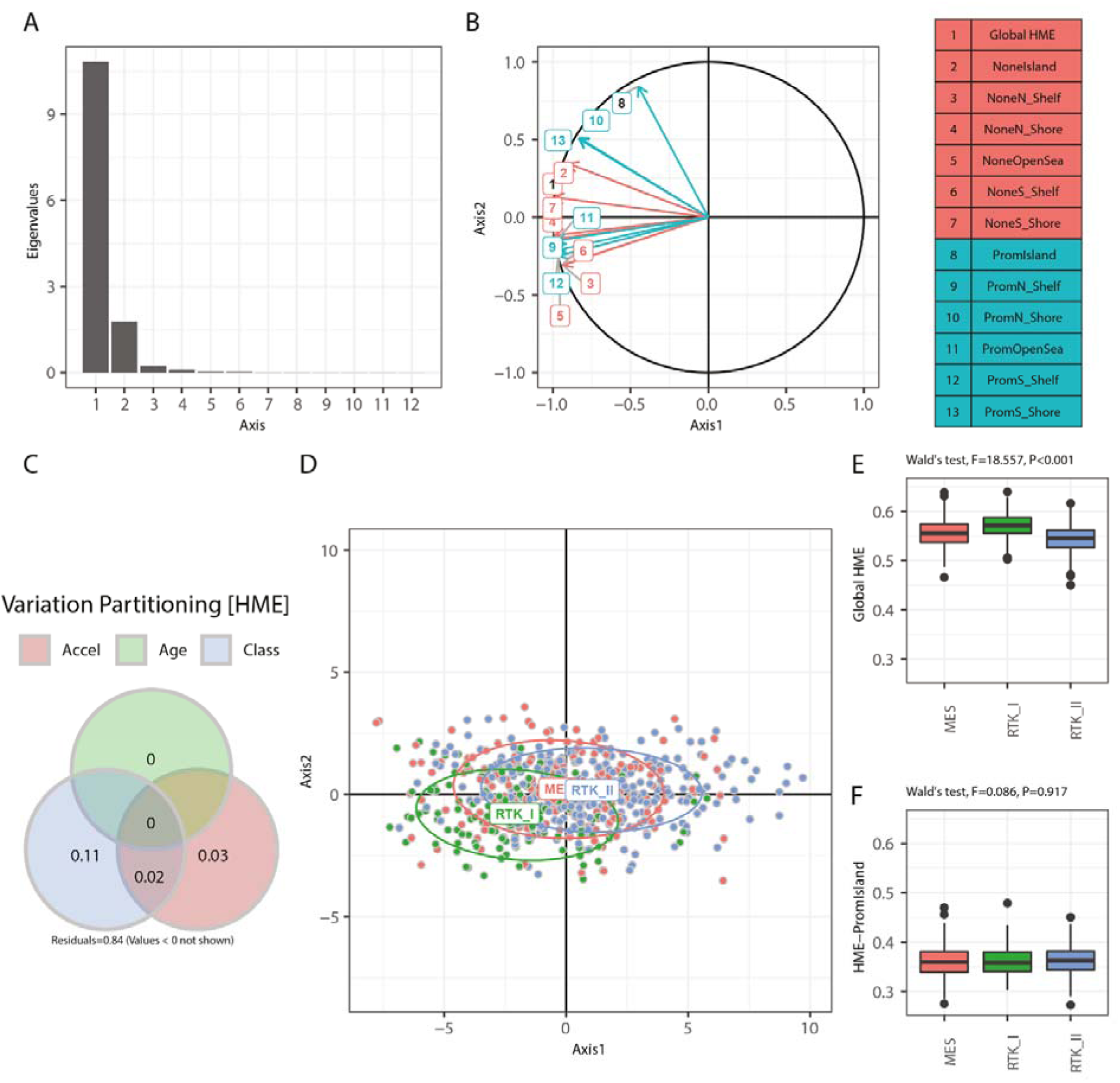
DNA methylation entropy (HME) of GBM in function of genome location. The entropy metrics based on DNA methylation (HME) was determined by genome location, constituted of the Island regions (CpG islands, shores, shelves or open sea) and promoter location (promoter or not in promoter). The Eigenvalues of the PCA of the dataset containing the HME metrics is visualized (A). The entropy (HME metrics) stratified by genome location was projected on the first vectorial plane of the PCA (B). The colors of the arrows and labels indicate the genome location in (green) or outside (red) of a promoter. (C) The contribution of DNAm age acceleration (Accel), age (Age) and GBM classification (Class) to HME was determined by variation partitioning and visualized in a Venn diagram containing the variation fractions. (D) The GBM samples are represented on the first vectorial plane of the PCA for global HME, annotated for GBM classification (MES, red; RTK I, green; RTK II, blue). Boxplot representation of global entropy (E) and entropy of CpGs located in promoter islands (F) is visualized by GBM classification. The association of classification with entropy by genome location was examined by Wald test (Additional File 1, Table S4).

This revealed some weak association of HM-entropy with GBM classification, explaining 13% (R2=0.131) of the variance, while age had very little impact (R2=0.005), and the contribution of DNAm age acceleration was also small (R2=0.050).

### DNA damage response (DDR) and DNAm age acceleration

Since all GBM patients, including the elderly, are treated with genotoxic therapy we had a closer look at the involvement of DNA damage response (DDR) genes in DNAm age acceleration. Evaluating only CpG probes associated with the promoter region of DDR genes as input (list as defined by Pearl et al., [39], 3947 CpGs, associated with 403 genes that might reveal age dependent treatment responses, did not yield any age related candidate CpGs.

DMPs related to DNAm age acceleration comprised 206 CpGs in 109 DDR genes whereof 22 CpGs in 8 genes were functional and comprised among others, the gene encoding an accessory subunit of DNA Polymerase Epsilon (*POLE4)*, the gene encoding Cyclin Dependent Kinase Inhibitor 2A *(CDKN2A), MGMT*, and the gene *Structural Maintenance of Chromosomes 1B (SMC1B)* (Supplementary Table S4). DMPs associated with tumor classification consisted of 272 CpGs in 138 DDR genes whereof 22 in 12 genes were functional (Supplementary Table S5). Three genes with functional CpGs were associated with both DNAm age acceleration and tumor classification and comprised *CDKN2A, MGMT*, and *SMC1B*. The methylation of the aforementioned functional CpGs identified in DDR genes is visualized in a heatmap (Supplementary, Figure S3).

### Age acceleration and outcome

Finally, we integrated DNAm age acceleration into the multivariable model for outcome for the two clinical trial cohorts, and included the following parameters: treatment (TMZ/RT→TMZ for EORTC/NCIC & LN-Pilot; TMZ for Nordic), MGMT status (*MGMT* methylation), and the interaction between treatment and *MGMT* methylation (predictive factor). DNAm age acceleration was significant in both cohorts, EORTC/NCIC & LN-Pilot p=0.0138 and Nordic p=0.00161 (Table 3). The predictive value of *MGMT* methylation (interaction between treatment and *MGMT* methylation) was confirmed in the EORTC/NCIC & LN-Pilot cohort (p<0.0001). The interaction term in the Nordic cohort was not significant (p=0.09). However, the number of *MGMT* methylated patients in the TMZ-arm was very small (n=13), suggesting lack of power for this test.

**Table 3:** Multivariable *Cox regression models including Age acceleration, GBM classification, global HM-entropy (HME) and HME at promoter CpGs.

| Model | Variables | EORTC/NCIC & LN-Pilot (N=177) |  |  |  | Nordic study (N=105) |  |  |  |
| --- | --- | --- | --- | --- | --- | --- | --- | --- | --- |
|  |  | modality | HR | z | Pr(>\|z\|) | modality | HR | z | Pr(>\|z\|) |
| Age acceleration [yrs] |  |  |  |  |  |  |  |  |  |
|  | Treatment (TRT) | TMZ+RT | 1.34862 | 1.31435 | 0.18873 | TMZ | 1.06845 | 0.23966 | 0.81059 |
|  | MGMT | MGMTmeth | 0.77273 | -1.10615 | 0.26866 | MGMTmeth | 1.13894 | 0.50912 | 0.61067 |
|  | Age Acceleration |  | 0.99124 | -2.46259 | **0.01379** |  | 0.98549 | -3.15493 | **0.00161** |
|  | TRT x MGMT | TMZ+RT x MGMTmeth | 0.27215 | -3.94628 | **0.00008** | TMZ x MGMTmeth | 0.47267 | -1.66716 | **0.09548** |
| GBM classification |  |  |  |  |  |  |  |  |  |
|  | Treatment (TRT) | TMZ+RT | 1.19859 | 0.80597 | 0.42026 | TMZ | 1.12226 | 0.41692 | 0.67674 |
|  | MGMT | MGMTmeth | 0.70503 | -1.52048 | 0.12839 | MGMTmeth | 1.05817 | 0.22379 | 0.82292 |
|  | GBM classification | GBM_RTK_I | 1.54086 | 1.91764 | **0.05516** | GBM_RTK_I | 1.12308 | 0.41675 | 0.67686 |
|  |  | GBM_RTK_II | 0.91674 | -0.48386 | 0.62848 | GBM_RTK_II | 0.52912 | -2.63579 | **0.00839** |
|  | TRT x MGMT | TMZ+RT x MGMTmeth | 0.31994 | -3.55697 | **0.00038** | TMZ x MGMTmeth | 0.53507 | -1.39736 | 0.16230 |
| Global HME |  |  |  |  |  |  |  |  |  |
|  | Treatment (TRT) | TMZ+RT | 1.12690 | 0.55400 | 0.57958 | TMZ | 1.08996 | 0.30807 | 0.75803 |
|  | MGMT | MGMTmeth | 0.69179 | -1.61442 | 0.10644 | MGMTmeth | 0.89983 | -0.42203 | 0.67300 |
|  | HM-Entropy |  | 3.18126 | 0.49623 | 0.61973 |  | 164.80788 | 1.64897 | **0.09915** |
|  |  | TMZ+RT x MGMTmeth | 0.34584 | -3.38245 | **0.00072** | TMZ x MGMTmeth | 0.54313 | -1.36378 | 0.17264 |
| HME promoter CpG |  |  |  |  |  |  |  |  |  |
|  | Treatment (TRT) | TMZ+RT | 1.15106 | 0.65024 | 0.51554 | TMZ | 1.17499 | 0.58063 | 0.56149 |
|  | MGMT | Methylated | 0.73235 | -1.33193 | 0.18288 | MGMTmeth d | 0.95937 | -0.16478 | 0.86912 |
|  | HM-Entropy |  | 0.04405 | -1.14097 | 0.25388 |  | 0.16488 | -0.58312 | 0.55982 |
|  | TRT x MGMT | TMZ+RT x MGMTmeth | 0.33173 | -3.48833 | **0.00049** | TMZ x MGMTmeth | 0.53779 | -1.39207 | 0.16390 |

\*Cox models, adjusted for Treatment (TRT), methylation of *MGMT* promoter, and the interaction of these two variables.

## Discussion

In this study, we investigated the DNA methylome of IDHwt GBM for age related differences that could hint at predictive or prognostic factors or indicate particular vulnerabilities for treatments in elderly patients. However, we found no strong direct associations, although we identified the methylation probe in *ELOVL2* that has reportedly the highest association with age as a single marker [5, 14] and *TRIM59* methylation, another robust marker for aging [32, 59]. *ELOVL2* is a GBM-relevant gene [15, 42]. However, methylation of this probe was not functional according to our analysis. *TRIM59* was among the genes associated with functional methylation. It encodes a protein with ubiquitin-transferase activity, and it has been associated with various regulatory processes and maybe involved in innate immune regulation.

We then used the metric of DNAm age and age acceleration proposed by the Horvath DNA methylation clock that has been trained on multiple tissues using the HM-27k array (does not comprise CpG probes *associated/annotated* with *ELOVL2*), and has been reported to be highly accurate to predict chronologic age (0.96). This methylation clock is considered independent of tissue type and mitotic potential [20, 22]. Various epigenetic clocks have been developed with potential as biomarkers. The aim is not only to determine accurate chronologic age, but to develop biological clocks for specific purposes, such as tissue specific sensors of disease, risk of disease, including cancer risk, or external stress, such as smoking history, or all-cause mortality (GrimAge) (reviewed in [5, 12, 22, 32]).

DNAm age acceleration observed for GBM averaged at almost 40 years, with a wide variability. No association was found with the patients’ sex. The affected gene sets comprised signatures associated with epigenetic features of neural precursor cells and specific neurotypes [25, 33, 34], suggestive of developmental features and cell of origin, but includes also cancer related signatures. The biological relevance is further supported by the fact that a subset of these signatures overlapped with those associated with functional methylation, implicating that the observed increased methylation was negatively associated with expression of the corresponding genes, affecting related pathways (Figure 3D, Supplementary Figure S2). It has been reported that CpGs whose methylation increases with age, are overrepresented near polycomb target genes that are important for stemness and cell differentiation and have been found frequently methylated in cancer [20, 50]. The investigation of mechanisms and biological consequences reflected in methylation aging clocks is an active field of research that has been extensively reviewed elsewhere [5, 12, 45].

When associating the observed DNAm age acceleration with outcome, we observed a significant effect, when analyzing the cohorts treated in clinical trials. Increased age acceleration in these IDHwt GBM was associated with better outcome, raising the question of the biological meaning of *DNAm age acceleration* in the context of these tumors. Therefore, we consider tumor related DNAm age acceleration as a measure for epigenetic distance associated with tumor development and GBM subtype. This is in accordance with the observation that DNAm age acceleration associated probes overlap largely with those associated with methylation–based GBM classification.

It has been suggested that deterioration of epigenetic maintenance contributes to age related changes [22], and more recently, it has been proposed that DNA break-induced epigenetic drift may contribute to aging [17]. Given the gross structural changes observed in GBM this is an interesting hypothesis. However, the numerous CNVs, characteristic of GBM, only explained a minor part (7%) of DNAm age of the tumors. Along the same lines, while a subset of clock probes was significantly associated with the methylation–based GBM classification, this was not dependent on their chromosomal location. Hence, this measure seems to comprehensively capture GBM related epigenetic changes that are associated in part with the GBM subclassification. The DNA methylation-based GBM classification is constituted of multidimensional information reflected by DNA methylation that are contributed by biological features such as purity, age acceleration, HM-entropy (variability of the DNA methylation), and other factors. The purity, reflecting infiltrating immune cells, measured by means of a DNA methylation variation, contributes to the differentiation of MES GBM samples, while DNAm age acceleration facilitates the differentiation of the RTK II GBM samples from those classified as RTK I and MES, and the HME discerns RTK I from the two others. Interestingly, among the significant pathways associated functional methylation and tumor classification, we identified previously reported expression-based GBM classification signatures [52]. This underlines the strong functional implication of DNA methylation on the expression phenotypes of the tumors and the coherence of the results.

The HM entropy metrics (12 distinct regions, and global entropy) exhibited genome-location dependent variation explained in part by the GBM subtypes (13%), but was basically not affected by age (0.5%). As an exception, HM entropy at promoter associated locations was lowest, and was not different across the GBM subtypes, indicating little permissiveness for variation in accordance with direct functional implications of methylated versus unmethylated status of promoters on gene expression.

Finally, we were interested in the DDR genes with functional methylation associated with DNAm age acceleration and classification that may yield mechanistic insights in the context of the genotoxic treatments that are part of the standard of care. Enhanced *POLE4* methylation (8 functional probes) was associated with DNAm age acceleration (Supplementary Figure S3). Deletion of *Pole-4* that encodes an accessory subunit of the DNA polymerase epsilon complex, has been reported to have no strong effects on its own in worms, however, in absence of the gene encoding the regulator of telomere elongation helicase 1 *(rtel-1)* apparently led to synthetic lethality due to impaired homologous recombination (HR) [6]. Similarly, six functional probes of *MGMT* were associated with DNAm age acceleration, whereof three were also associated with GBM subclassification. Functional probes in *CDKN2A* and *SMCB1* were also associated with both. Other functional probes were only associated with tumor classification, e.g. the gene encoding the Fanconi anemia complementation group M protein *(FANCM)* or the gene encoding the AlkB Homolog 1, Histone H2A Dioxygenase *(ALKBH1)*. The former is involved in homology directed DNA repair, and the latter takes part in the repair of DNA alkylation damage and has recently been associated with the regulation of the level of N^6^-methyladenine (N^6^-mA) DNA modifications, implicated in epigenetic regulation of gene expression relevant in GBM [57]. While *MGMT* methylation is a known predictive factor for responsiveness to the alkylating agent TMZ in GBM, it remains to be explored, whether any of the other identified probes and their associated genes indicate potentially actionable vulnerabilities.

In conclusion, once removing all high grade gliomas not classified as GBM IDHwt WHO grade 4, the spectrum of the tumors seems to be similar across adult age, at least from a DNA methylation point of view. DNAm differences of the tumors quantified as age acceleration or HME contribute to features of tumor subtypes, while age is hardly reflected. Interestingly, the epigenetic distance measured as DNAm age acceleration was associated with better outcome in the cohorts treated in clinical trials also for elderly GBM patients.

## Data Availability

All data will be accessible at the Gene Expression Omnibus - NCBI, http://www.ncbi.nlm.nih.gov/geo/

https://www.ncbi.nlm.nih.gov/geo/query/acc.cgi?acc=GSE195684

https://www.ncbi.nlm.nih.gov/geo/query/acc.cgi?acc=GSE195640

https://www.ncbi.nlm.nih.gov/geo/query/acc.cgi?acc=GSE60274

https://www.ncbi.nlm.nih.gov/geo/query/acc.cgi?acc=GSE104293

http://cancergenome.nih.gov

https://www.ncbi.nlm.nih.gov/geo/query/acc.cgi?acc=GSE109381

## Acknowledgements

We thank the patients and their families for their support and participation, and the treating clinical centers for collaboration. The results published here are in part based upon data generated by The Cancer TCGA Genome Atlas pilot project established by the NCI and NHGRI. Information about TCGA and the investigators and institutions WHO constitute the TCGA research network can be found at “http://cancergenome.nih.gov”. The dbGaP accession number to the specific version of the TCGA data set is phs000178.v8.p7.

## Author’s contributions

M.E.H. conceived and designed the study together with A.M. and R.S.; P.B. designed, and performed the biostatistical analyses, and interpretation; C.M., R.S., M.W., B.H.G., H.S., A.M., M.T., J.M.M.G. and M.v.B. were the main recruiters for the clinical trials and provided respective study material that made this project possible. A.v.D. provided the methylation-based classification of the datasets. M.E.H. collected data and material, coordinated sample analysis, interpreted the results and wrote the manuscript with P.B. All authors read and approved the final manuscript.

## Ethics approval and consent to participate

Patients from the clinical trials cohorts provided written informed consent for translational research in the context of their enrollment into the clinical trials. The Lausanne Pilot trial was approved by the Commission cantonale (Vaud) d’éthique de la recherche sur l’être humain (CER-VD), Lausanne, Switzerland and the Commission Cantonale d’Ethique de la Recherche sur l’être humain(CCER), Geneva, Switzerland. The EORTC 26981/NCIC CE.3 trial (trial registration number NCT00006353) was approved by the ethics committees of all 85 participating centers, ethical approval from the lead center was approved by the Commission cantonale (Vaud) d’éthique de la recherche sur l’être humain (CER-VD), Lausanne, Switzerland. The study was conducted according to GCP under the auspices/responsibility and sponsorship of the European Organisation for Research and Treatment of Cancer (EORTC, Brussels, Belgium). The trial was audited by the American Food and Drug Administration (FDA) with no major findings reported. The Nordic trial (trial registration number ISRCTN81470623) received ethics approval from the Linköping regional ethics committee, Linköping, Sweden. The study has been performed according to institutional and international guidelines and regulations as previously reported [31, 46, 48].

