## Supplementary Information for "DNA methylation-based age acceleration observed in IDH wild-type glioblastoma is associated with better outcome - including in elderly patients"

**Supplementary File**

**Supplementary Tables S1-S5**

- **Table S1.** Description of full EORTC/NCIC & LN-Pilot and Nordic datasets.
- **Table S2.** DNA methylation associated with patient age.
- **Table S3.** Associations of methylation based entropy by genomic region and GBM classification.
- **Table S4.** Description of the CpGs located on DDR genes, associated with Age Acceleration and considered as functional.
- **Table S5.** Functional CpGs located on DDR genes, associated with GBM classification.

**Supplementary Figures S1-S3**

- **Figure S1.** Methylation-based classification of tumors of all patients from EORTC/NCIC & LN-Pilot and Nordic studies.
- **Figure S2.** Pathways associated with functional methylation related to DNAm age acceleration and GBM classification.
- **Figure S3.** Heatmap of DNA methylation of functional CpGs in DDR genes significantly associated with DNAm age acceleration and GBM classification.

**Table S1:** Description of full EORTC/NCIC & LN-Pilot and Nordic datasets

| | **EORTC/NCIC & LN-Pilot (N = 219)** | **Nordic (N = 116)** |
|---|---|---|
| **age** | | |
| minimum | 25.00 | 60.05 |
| median (IQR) | 53.50 (48.00, 60.00) | 70.41 (66.51, 74.09) |
| mean (sd) | 52.86 ± 9.77 | 70.36 ± 4.80 |
| maximum | 70.00 | 83.06 |
| Unknown/Missing | 1 (0.46%) | 0 (0.00%) |
| **DNAage** | | |
| minimum | 31.18 | 51.90 |
| median (IQR) | 90.12 (71.47, 108.11) | 103.19 (83.53, 123.92) |
| mean (sd) | 91.26 ± 27.23 | 105.47 ± 25.88 |
| maximum | 167.65 | 180.46 |
| **Accel1** | | |
| minimum | -24.40 | -12.07 |
| median (IQR) | 35.66 (21.11, 54.72) | 32.24 (15.48, 52.18) |
| mean (sd) | 38.37 ± 26.03 | 35.11 ± 25.26 |
| maximum | 107.34 | 108.46 |
| Unknown/Missing | 1 (0.46%) | 0 (0.00%) |
| **Sex** | | |
| F | 78 (36) | 43 (37) |
| M | 141 (64) | 73 (63) |
| **Performance Score** | | |
| 0 | 95 (44) | 29 (25) |
| 1 | 107 (49) | 61 (53) |
| 2 | 16 (7) | 22 (19) |
| 3 | 0 (0) | 4 (3) |
| Unknown/Missing | 1 (0.46%) | 0 (0.00%) |
| **MGMTscore** | | |
| minimum | -10.48 | -4.09 |
| median (IQR) | -0.73 (-2.68, 3.80) | -0.93 (-2.66, 3.70) |
| mean (sd) | 0.29 ± 3.40 | 0.31 ± 3.33 |
| maximum | 7.35 | 8.16 |
| **MGMTclass** | | |
| M | 107 (49) | 56 (48) |
| U | 112 (51) | 60 (52) |
| **hCIMP** | | |
| cimp | 15 (7) | 1 (1) |
| noncimp | 199 (93) | 115 (99) |
| Unknown/Missing | 5 (2.28%) | 0 (0.00%) |

Continued next page

Table S1, continued from previous page

| | EORTC/NCIC & LN-Pilot (N = 219) | Nordic (N = 116) |
|---|---|---|
| **NMP classification** | | |
| A_IDH | 2 (1) | 0 (0) |
| A_IDH_HG | 11 (5) | 1 (1) |
| ANA_PA | 1 (0) | 0 (0) |
| CONTR_HEMI | 1 (0) | 0 (0) |
| CONTR_INFLAM | 3 (1) | 7 (6) |
| CONTR_REACT | 1 (0) | 0 (0) |
| CONTR_WM | 1 (0) | 0 (0) |
| DMG_K27 | 1 (0) | 0 (0) |
| EPN_RELA | 1 (0) | 0 (0) |
| GBM_G34 | 1 (0) | 0 (0) |
| GBM_MES | 69 (32) | 43 (37) |
| GBM_MID | 2 (1) | 2 (2) |
| GBM_MYCN | 1 (0) | 0 (0) |
| GBM_RTK_I | 32 (15) | 20 (17) |
| GBM_RTK_II | 85 (39) | 42 (36) |
| O_IDH | 2 (1) | 0 (0) |
| PLEX_PED_B | 2 (1) | 1 (1) |
| PXA | 3 (1) | 0 (0) |
| **Study** | | |
| EORTC/NCIC & LN-Pilot | 219 (100) | 0 (0) |
| Nordic | 0 (0) | 116 (100) |

**Table S2**. DNA methylation associated with patient age

| ProbeID | symbol | slope (b) | R-squared | F-value | Pr(>F) | P-Bonferroni | Location type | Relation Island | *Element Type | Accel | Class | Corr (Expr Methyl) | p-value | P-Bonf |
|---|---|---|---|---|---|---|---|---|---|---|---|---|---|---|
| cg03514351 | LEPR | 0.044365 | 0.05471527 | 34.3242168 | 7.7391E-09 | 0.00279959 | gene; prom | Island | Prom/Enh | **TRUE** | **TRUE** | | | |
| cg08816037 | IFT80; TRIM59 | 0.03865695 | 0.05110595 | 31.9380543 | 2.4741E-08 | 0.00894991 | promoter | Island | Prom/Enh | TRUE | FALSE | -0.56471 | 0.000511 | 0.028469 |
| cg16867657 | ELOVL2-AS1; ELOVL2 | 0.0238864 | 0.05436931 | 34.0947058 | 8.6519E-09 | 0.0031298 | promoter | Island | Prom/Enh | **TRUE** | **TRUE** | | | |
| cg06335867 | NXPH1 | 0.04966675 | 0.06025499 | 38.0222419 | 1.2941E-09 | 0.00046815 | gene | Island | | **TRUE** | **TRUE** | | | |
| cg12597389 | NXPH1 | 0.04007597 | 0.05428946 | 34.0417571 | 8.8775E-09 | 0.00321137 | gene | Island | | TRUE | FALSE | | | |
| cg12744812 | MNX1-AS1 | 0.02823639 | 0.04488323 | 27.8664939 | 1.8264E-07 | 0.06606765 | extend | Island | | FALSE | TRUE | | | |
| cg16197925 | FEZF1-AS1; FEZF1 | 0.0279542 | 0.04686293 | 29.1560574 | 9.6744E-08 | 0.03499674 | promoter | Island | Prom/Enh | TRUE | FALSE | | | |
| cg26170604 | NXPH1 | 0.04976372 | 0.04570145 | 28.3988245 | 1.4046E-07 | 0.05081095 | gene | Island | Enhancer | FALSE | TRUE | | | |
| cg26818735 | TWIST1 | 0.05287926 | 0.04393627 | 27.2515378 | 2.4747E-07 | 0.08952102 | promoter | Island | Prom/Enh | TRUE | FALSE | -0.60784 | 0.000147 | 0.013656 |
| cg03323636 | NRIP3 | 0.04335599 | 0.05311561 | 33.2644194 | 1.2957E-08 | 0.00468715 | promoter | Island | Prom/Enh | TRUE | FALSE | -0.59272 | 0.000232 | 0.017835 |
| cg23132624 | KL | 0.04874967 | 0.0545758 | 34.2316671 | 8.095E-09 | 0.00292831 | promoter | Island | Prom/Enh | TRUE | FALSE | | | |
| cg02970384 | ACTA1 | 0.02676012 | 0.05000415 | 31.2132523 | 3.5262E-08 | 0.01275585 | gene | Island | Prom/Enh | TRUE | FALSE | | | |
| cg06527052 | TMEM167B | 0.00617915 | 0.04850109 | 30.2271997 | 5.7163E-08 | 0.02067838 | promoter | S_Shore | Prom/Enh | FALSE | FALSE | | | |
| cg23704082 | IFT80; TRIM59 | 0.04009039 | 0.04700147 | 29.2464994 | 9.2535E-08 | 0.03347408 | promoter | Island | Prom/Enh | TRUE | FALSE | | | |
| cg06470822 | | 0.03211127 | 0.0456596 | 28.3715794 | 1.4236E-07 | 0.05149791 | intergenic | Island | | **TRUE** | **TRUE** | | | |
| cg20974724 | FEZF1-AS1; FEZF1 | 0.03674432 | 0.0519421 | 32.4892225 | 1.8905E-08 | 0.00683876 | promoter | Island | Prom/Enh | TRUE | FALSE | | | |
| cg25098077 | PDLIM1 | 0.03457268 | 0.04716098 | 29.3506692 | 8.7915E-08 | 0.03180264 | promoter | Island | Prom/Enh | TRUE | FALSE | | | |
| cg01994205 | RNF219-AS1;POU4F1 | 0.04504875 | 0.04792986 | 29.8532717 | 6.8677E-08 | 0.0248437 | gene; prom | Island | Prom/Enh | TRUE | FALSE | | | |
| cg02796545 | KL | 0.0353594 | 0.05429518 | 34.0455517 | 8.8611E-09 | 0.00320546 | promoter | Island | Prom/Enh | TRUE | FALSE | | | |

*genehancer 2.0

**Table S3.** Associations of methylation based entropy by genomic region and GBM classification.

| Characteristic | Prom | CpG-Island relationship | MES mean (sd) | RTK I mean (sd) | RTK II mean (sd) | GBM classification | | Study | | GBM classification x Study | |
|---|---|---|---|---|---|---|---|---|---|---|---|
| | | | | | | F-value | Pr(>F) | F-value | Pr(>F) | F-value | Pr(>F) |
| NoneIsland | no | CpGs-Island | 0.557 (0.029) | 0.567 (0.029) | 0.55 (0.03) | 6.207053 | 0.00214944 | 0.05208871 | 0.984304 | 0.2322369 | 0.966069364 |
| NoneN_Shelf | no | N_Shelf | 0.639 (0.043) | 0.685 (0.038) | 0.629 (0.042) | 39.4388337 | 8.4277E-17 | 0.30650644 | 0.8207 | 1.2374211 | 0.285187616 |
| NoneN_Shore | no | N_shore | 0.653 (0.034) | 0.675 (0.032) | 0.638 (0.035) | 24.9416396 | 4.0024E-11 | 0.12473897 | 0.945475 | 0.7216483 | 0.632292687 |
| NoneOpenSea | no | Open Sea | 0.643 (0.039) | 0.673 (0.036) | 0.625 (0.039) | 30.9767708 | 1.6161E-13 | 0.20681642 | 0.891689 | 0.9450378 | 0.462079214 |
| NoneS_Shelf | no | S_Shelf | 0.637 (0.043) | 0.683 (0.038) | 0.627 (0.042) | 38.2902463 | 2.3248E-16 | 0.29527014 | 0.82883 | 1.1771564 | 0.316748706 |
| NoneS_Shore | no | S_shore | 0.64 (0.034) | 0.66 (0.032) | 0.624 (0.034) | 23.592329 | 1.3922E-10 | 0.11733844 | 0.949928 | 0.5937028 | 0.735497569 |
| PromIsland | yes | CpGs-Island | 0.362 (0.032) | 0.361 (0.03) | 0.363 (0.029) | 0.08626142 | 0.91736597 | 0.14667742 | 0.93181 | 1.8152771 | 0.093836959 |
| PromN_Shelf | yes | N_Shelf | 0.662 (0.035) | 0.686 (0.033) | 0.644 (0.036) | 28.3923689 | 1.6908E-12 | 0.13803558 | 0.93727 | 1.1699905 | 0.320671299 |
| PromN_Shore | yes | N_shore | 0.55 (0.026) | 0.545 (0.027) | 0.535 (0.025) | 6.71057346 | 0.00131324 | 0.04780861 | 0.986145 | 0.3061348 | 0.933792612 |
| PromOpenSea | yes | Open Sea | 0.648 (0.034) | 0.666 (0.032) | 0.63 (0.034) | 22.4423925 | 4.0449E-10 | 0.11010916 | 0.954191 | 0.984101 | 0.435052346 |
| PromS_Shelf | yes | S_Shelf | 0.639 (0.037) | 0.67 (0.033) | 0.622 (0.037) | 35.6277052 | 2.4762E-15 | 0.20068678 | 0.895914 | 1.1349293 | 0.340387478 |
| PromS_Shore | yes | S_shore | 0.548 (0.026) | 0.543 (0.027) | 0.532 (0.025) | 7.39443155 | 0.00067343 | 0.05563908 | 0.982727 | 0.27667 | 0.947916261 |
| Global HME | no | none | 0.556 (0.029) | 0.571 (0.027) | 0.545 (0.029) | 18.5570792 | 1.5293E-08 | 0.02850008 | 0.993511 | 0.3939696 | 0.883032642 |

Association tests with GBM classification, study origin and the interaction between these both variables for the entropy metrics based on DNA methylation Entropy (HME) and stratified by the Island regions (CpG islands, shores, shelves or open sea) and promoter location status (promoter or not in promoter). The linear models are compared by Wald's test using the sandwich covariance matrix (type HC3) and F-statistic to compensate heteroskedasticity.

**Table S4**: Description of the CpGs located on DDR genes, associated with Age Acceleration and considered as functional.

| ProbeID | entrezID | Symbol | RnaseqID | Correlation Expr vs Methyl | | | Age | | Age acceleration | | Classification |
|---|---|---|---|---|---|---|---|---|---|---|---|
| | | | | cor | p.value | padj | slope | R2 | slope | R2 | R2 |
| cg05382305 | 164045 | HFM1 | HFM1\|164045 | -0.4857 | 0.0034 | 0.0938 | -0.0027 | 0.0010 | 0.0082 | 0.0417 | 0.0035 |
| cg23032045 | 164045 | HFM1 | HFM1\|164045 | -0.5440 | 0.0009 | 0.0479 | -0.0123 | 0.0090 | 0.0128 | 0.0462 | 0.0119 |
| cg15091337 | 56655 | POLE4 | POLE4\|56655 | -0.6751 | 0.0000 | 0.0034 | 0.0019 | 0.0002 | 0.0163 | 0.0780 | 0.0216 |
| cg12290764 | 56655 | POLE4 | POLE4\|56655 | -0.7347 | 0.0000 | 0.0009 | 0.0015 | 0.0001 | 0.0243 | 0.1361 | 0.0253 |
| cg02058002 | 56655 | POLE4 | POLE4\|56655 | -0.7387 | 0.0000 | 0.0009 | 0.0001 | 0.0000 | 0.0243 | 0.1157 | 0.0198 |
| cg02307033 | 56655 | POLE4 | POLE4\|56655 | -0.7894 | 0.0000 | 0.0007 | 0.0036 | 0.0005 | 0.0267 | 0.1150 | 0.0270 |
| cg20919922 | 56655 | POLE4 | POLE4\|56655 | -0.7459 | 0.0000 | 0.0009 | 0.0039 | 0.0003 | 0.0348 | 0.1180 | 0.0260 |
| cg12696259 | 56655 | POLE4 | POLE4\|56655 | -0.6499 | 0.0000 | 0.0065 | 0.0178 | 0.0088 | 0.0314 | 0.1285 | 0.0224 |
| cg13690354 | 56655 | POLE4 | POLE4\|56655 | -0.7045 | 0.0000 | 0.0017 | 0.0098 | 0.0027 | 0.0283 | 0.1042 | 0.0347 |
| cg05778415 | 56655 | POLE4 | POLE4\|56655 | -0.6120 | 0.0001 | 0.0153 | 0.0078 | 0.0021 | 0.0288 | 0.1352 | 0.0151 |
| cg20142358 | 57599 | WDR48 | WDR48\|57599 | -0.4908 | 0.0031 | 0.0882 | 0.0032 | 0.0041 | 0.0045 | 0.0389 | 0.0181 |
| cg15438497 | 6596 | HLTF | HLTF\|6596 | -0.4812 | 0.0038 | 0.0982 | -0.0028 | 0.0008 | 0.0139 | 0.0956 | 0.0515 |
| cg03678609 | 6596 | HLTF | HLTF\|6596 | -0.4980 | 0.0026 | 0.0789 | -0.0025 | 0.0004 | 0.0212 | 0.1229 | 0.0827 |
| cg07562918 | 1029 | CDKN2A | CDKN2A\|1029 | -0.6429 | 0.0000 | 0.0076 | 0.0043 | 0.0038 | 0.0096 | 0.0873 | 0.1398 |
| cg14194875 | 4255 | MGMT | MGMT\|4255 | -0.7151 | 0.0000 | 0.0014 | 0.0188 | 0.0107 | 0.0160 | 0.0365 | 0.0429 |
| cg00618725 | 4255 | MGMT | MGMT\|4255 | -0.6258 | 0.0001 | 0.0113 | 0.0213 | 0.0165 | 0.0145 | 0.0364 | 0.0373 |
| cg12434587 | 4255 | MGMT | MGMT\|4255 | -0.7983 | 0.0000 | 0.0007 | 0.0178 | 0.0052 | 0.0257 | 0.0513 | 0.0188 |
| cg02802904 | 4255 | MGMT | MGMT\|4255 | -0.5109 | 0.0020 | 0.0701 | 0.0148 | 0.0099 | 0.0128 | 0.0345 | 0.0165 |
| cg12981137 | 4255 | MGMT | MGMT\|4255 | -0.7269 | 0.0000 | 0.0010 | 0.0271 | 0.0120 | 0.0235 | 0.0429 | 0.0415 |
| cg02941816 | 4255 | MGMT | MGMT\|4255 | -0.6880 | 0.0000 | 0.0025 | 0.0171 | 0.0114 | 0.0132 | 0.0324 | 0.0282 |
| cg20808578 | 1454 | CSNK1E | CSNK1E\|1454 | -0.5711 | 0.0004 | 0.0323 | 0.0011 | 0.0002 | 0.0064 | 0.0331 | 0.0001 |
| cg22884516 | 27127 | SMC1B | SMC1B\|27127 | -0.5134 | 0.0018 | 0.0687 | 0.0212 | 0.0117 | 0.0313 | 0.1193 | 0.0517 |

**Table S5.** Functional CpGs located on DDR genes, associated with GBM classification.

| ProbeID | entrezID | Symbol | RnaseqID | Correlation Expr vs Methyl | | | Age | | Age acceleration | | GBM classification |
|---|---|---|---|---|---|---|---|---|---|---|---|
| | | | | corr | p.value | padj | slope | R2 | slope | R2 | R2 |
| cg15438497 | 6596 | HLTF | HLTF\|6596 | -0.4812 | 0.0038 | 0.0982 | -0.0028 | 0.0008 | 0.0139 | 0.0956 | 0.0515 |
| cg03678609 | 6596 | HLTF | HLTF\|6596 | -0.4980 | 0.0026 | 0.0789 | -0.0025 | 0.0004 | 0.0212 | 0.1229 | 0.0827 |
| cg24183261 | 2138 | EYA1 | EYA1\|2138 | -0.5165 | 0.0017 | 0.0662 | -0.0004 | 0.0000 | 0.0023 | 0.0057 | 0.0456 |
| cg13043862 | 2138 | EYA1 | EYA1\|2138 | -0.5174 | 0.0017 | 0.0662 | 0.0026 | 0.0018 | 0.0022 | 0.0061 | 0.0389 |
| cg13601799 | 1029 | CDKN2A | CDKN2A\|1029 | -0.7384 | 0.0000 | 0.0009 | 0.0086 | 0.0116 | 0.0024 | 0.0042 | 0.0831 |
| cg03079681 | 1029 | CDKN2A | CDKN2A\|1029 | -0.6958 | 0.0000 | 0.0020 | 0.0059 | 0.0102 | 0.0014 | 0.0026 | 0.0927 |
| cg07562918 | 1029 | CDKN2A | CDKN2A\|1029 | -0.6429 | 0.0000 | 0.0076 | 0.0043 | 0.0038 | 0.0096 | 0.0873 | 0.1398 |
| cg10848754 | 1029 | CDKN2A | CDKN2A\|1029 | -0.7062 | 0.0000 | 0.0017 | 0.0125 | 0.0208 | 0.0061 | 0.0231 | 0.0976 |
| cg14430974 | 1029 | CDKN2A | CDKN2A\|1029 | -0.7017 | 0.0000 | 0.0017 | 0.0143 | 0.0243 | 0.0067 | 0.0252 | 0.0969 |
| cg14194875 | 4255 | MGMT | MGMT\|4255 | -0.7151 | 0.0000 | 0.0014 | 0.0188 | 0.0107 | 0.0160 | 0.0365 | 0.0429 |
| cg00618725 | 4255 | MGMT | MGMT\|4255 | -0.6258 | 0.0001 | 0.0113 | 0.0213 | 0.0165 | 0.0145 | 0.0364 | 0.0373 |
| cg12981137 | 4255 | MGMT | MGMT\|4255 | -0.7269 | 0.0000 | 0.0010 | 0.0271 | 0.0120 | 0.0235 | 0.0429 | 0.0415 |
| cg25419628 | 9937 | DCLRE1A | DCLRE1A\|9937 | -0.5535 | 0.0007 | 0.0432 | 0.0069 | 0.0138 | 0.0042 | 0.0248 | 0.0591 |
| cg03727700 | 9937 | DCLRE1A | DCLRE1A\|9937 | -0.5473 | 0.0008 | 0.0470 | 0.0027 | 0.0043 | 0.0030 | 0.0248 | 0.0429 |
| cg18787244 | 9937 | DCLRE1A | DCLRE1A\|9937 | -0.5947 | 0.0002 | 0.0215 | 0.0021 | 0.0033 | 0.0020 | 0.0140 | 0.0443 |
| cg03817911 | 493861 | EID3 | EID3\|493861 | -0.5042 | 0.0023 | 0.0743 | 0.0019 | 0.0002 | 0.0001 | 0.0000 | 0.1450 |
| cg09096528 | 1019 | CDK4 | CDK4\|1019 | -0.6812 | 0.0000 | 0.0030 | 0.0036 | 0.0059 | 0.0000 | 0.0000 | 0.0424 |
| cg11869215 | 328 | APEX1 | APEX1\|328 | -0.5535 | 0.0007 | 0.0432 | 0.0042 | 0.0076 | -0.0025 | 0.0127 | 0.0428 |
| cg06755612 | 57697 | FANCM | FANCM\|57697 | -0.4958 | 0.0028 | 0.0795 | 0.0008 | 0.0003 | -0.0008 | 0.0015 | 0.0353 |
| cg25564554 | 8846 | ALKBH1 | ALKBH1\|8846 | -0.5120 | 0.0019 | 0.0690 | 0.0043 | 0.0097 | -0.0013 | 0.0039 | 0.0433 |
| cg13367381 | 10054 | UBA2 | UBA2\|10054 | -0.5521 | 0.0007 | 0.0432 | 0.0022 | 0.0050 | -0.0013 | 0.0087 | 0.0359 |
| cg22884516 | 27127 | SMC1B | SMC1B\|27127 | -0.5134 | 0.0018 | 0.0687 | 0.0212 | 0.0117 | 0.0313 | 0.1193 | 0.0517 |

### Supplementary Figures

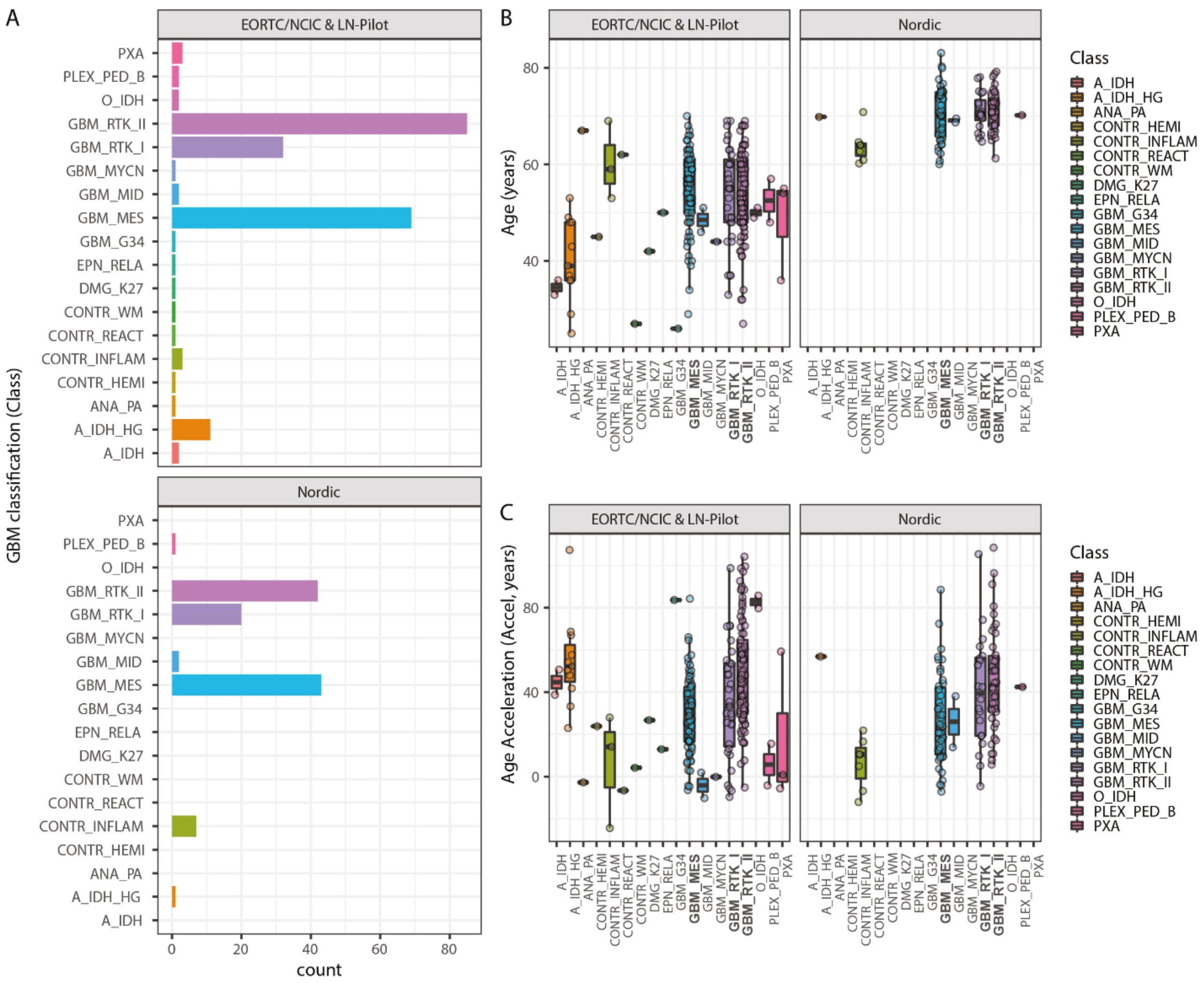


**Figure S1.** Methylation-based classification of tumors of all patients from EORTC/NCIC & LN-Pilot and Nordic studies. (A) The number of patients by methylation-based classification subtype is visualized for patients in the EORTC/NCIC & LN-Pilot and Nordic studies. The variables age (B) and DNAm age acceleration (C) are illustrated in function of the molecular subgroups and the study origin.

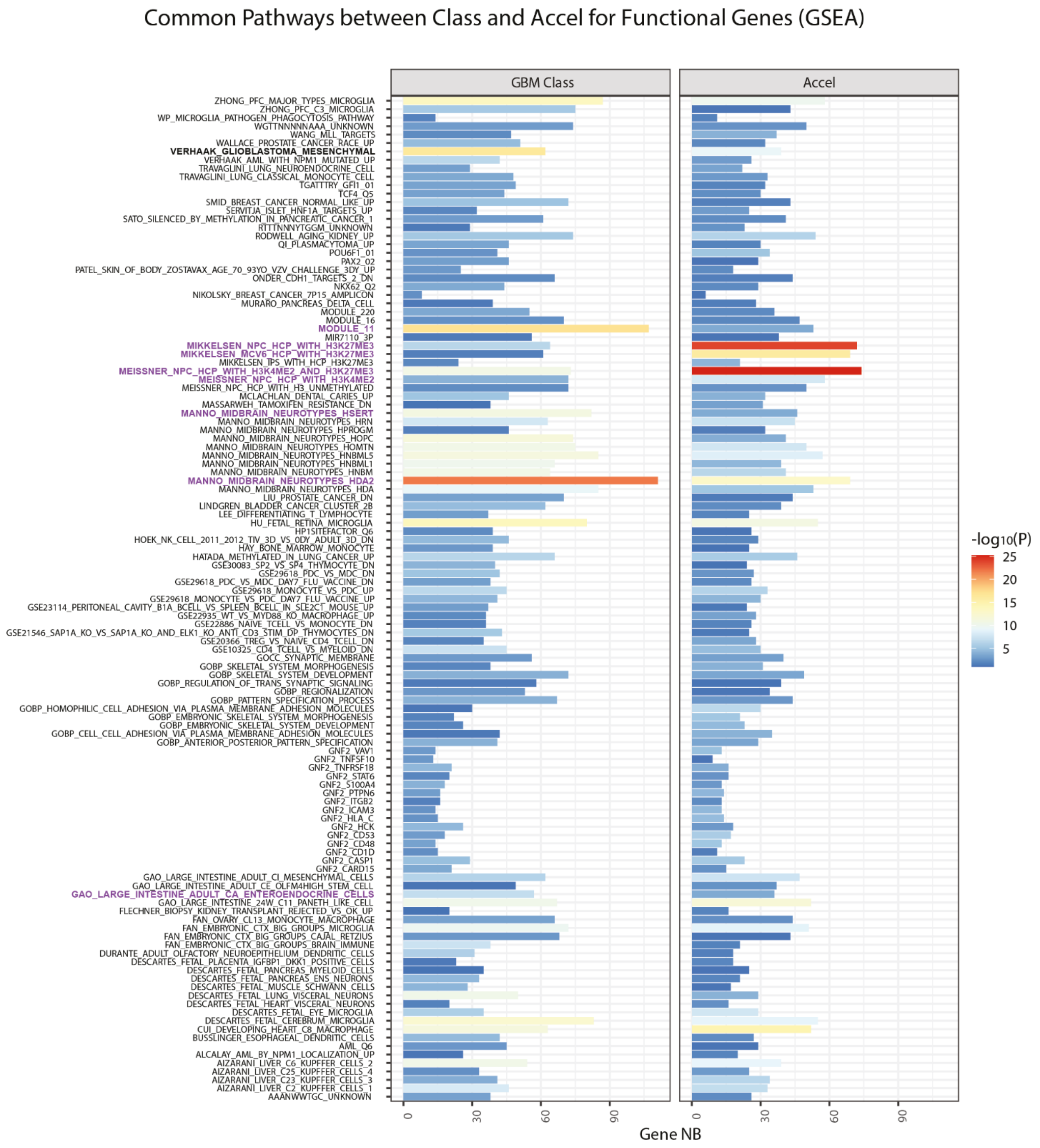


**Figure S2.** Pathways associated with functional methylation related to DNAm age acceleration and GBM classification. Gene set enrichment analysis (GSEA) was established for functional candidate CpG-probes located in gene promoters associated with age, DNAm age acceleration, and GBM classification. Functional methylated positions were defined as functional, when the correlation coefficient was inferior to -0.3 (negative effect of methylation on gene expression) and the q-value was less than 0.1. No significant “functional” pathways were detected for age. The “functional” pathways associated with DNAm age accel (n=167) mostly (119, 71%) overlapped with those associated with classification (n=294). The list retained, comprises selected pathways significantly enriched for both, DNAm age acceleration and GBM classification. The number (NB) of genes per gene set is indicated and the p-value is represented by the color code. The gene sets overlapping with those identified significant for genome wide DNA methylation (Figure 4D) for classification and DNAm age acceleration are highlighted in color. Of note, the Verhaak signature for mesenchymal GBM is part of this list, marked in bold. The pathways are listed in alphabetical order. GSEA investigation is based on the MSigDB database.

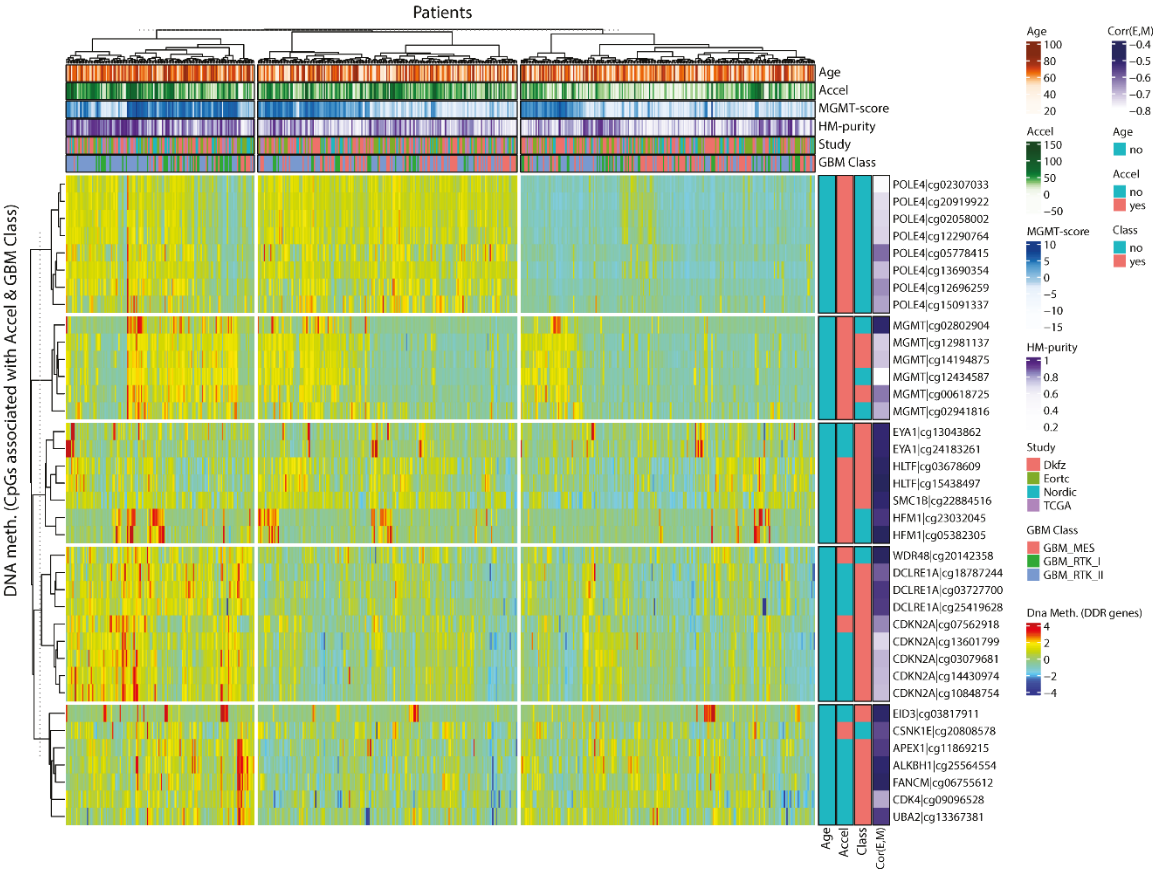


**Figure S3.** Heatmap of DNA methylation of functional CpGs in DDR genes significantly associated with DNAm age acceleration and GBM classification. The clustering was obtained by Ward's algorithm using Euclidean distance based on centered and scaled data. The samples and the methylation of the functional CpGs associated with DDR genes were classified into 3 clusters (consensus k-means clustering for 200 repetitions) and 5 clusters (consensus k-means clustering for 200 repetitions), respectively. The samples are annotated with age (age in years), DNAm age acceleration (Accel1, in years), *MGMT* methylation score (MGMTscore), Purity index based on DNA methylation (HMpurity), study origin (study) and GBM classification (Class). The probes are annotated for statistical significance associated with age (Age), DNAm age acceleration (Accel) or GBM classification (Class), yes, red; no, blue. The Spearman's correlation of methylation with expression (measure of functionality) is given for each CpG (corr).
